# Assessment of the Ten Steps to Successful Breastfeeding of the Baby-Friendly Hospital Initiative at Healthcare Facilities in East Java, Indonesia

**DOI:** 10.1101/2022.07.26.22278047

**Authors:** Pande Putu Januraga, Ngakan Putu Anom Harjana, Mellysa Kowara, Putu Ayu Indrayathi, Dinar SM Lubis, I Gusti Ngurah Edi Putra, Doddy Izwardi

## Abstract

**Background:** The implementation of the Ten Steps to Successful Breastfeeding (TSSB) of the Baby-Friendly Hospital Initiative (BFHI) has been shown to be associated with improved breastfeeding outcomes. Indonesia still lags other countries in promoting breastfeeding in the first hour and 6 months of exclusive breastfeeding. This study aimed to assess health facilities’ compliance with the TSSB in Indonesia.

**Methods:** A cross-sectional survey of 242 health facility managers and 130 postpartum mothers across hospitals, community health centers (puskesmas), and private clinics at 5 sites in the East Java province was conducted between January and June, 2019. The health facility managers and mothers were interviewed using questionnaires consisting of questions adopted from the TSSB. The data were analyzed using the descriptive method to present the level of compliance with each step, the overall steps, comparing self-appraisal and validating methods, and comparing results by study site and type of health facility.

**Results:** The assessment data showed various levels of compliance with the TSSB, with scores ranging from 65.47 to 98.09 (mean score 77.5 on a scale from 0 to 100). The highest compliance was found in step 8 (breastfeeding on demand) and the lowest in step 7 (rooming-in). The validation results showed a significantly lower compliance with steps 3, 4, 8, and 9 compared with the self-appraisal. The assessment of compliance with the TSSB also showed certain variations between the site and the type of health facility.

**Conclusion:** We recommend that policy makers and managers of health facilities formulate effective and appropriate policies to increase institutional compliance with the TSSB. Greater efforts are needed to facilitate puskesmas and private clinics in implementing the TSSB for better breastfeeding outcomes.

## Introduction

The recent evaluation of exclusive breastfeeding across 194 nations in the Global Breastfeeding report found that in 2013–2018, 43% of newborns initiated breastfeeding within 1 hour of birth, with only 41% of infants under 6 months of age receiving exclusive breastfeeding [1]. These figures are far from the collective global rates targeted for 2030, which are 70% for initiating breastfeeding in the first hour and 70% for exclusive breastfeeding. Breastfeeding initiation in the first hour after birth and exclusive breastfeeding are known to have significant short-term benefits for children’s health, mainly protection against morbidity and mortality from infectious diseases [2]. Moreover, the long-term effects of exclusive breastfeeding on blood pressure, diabetes, and related indicators, serum cholesterol, overweight and obesity, and intellectual performance have been documented in a systematic review and meta-analysis [3]. Therefore, efforts to improve the coverage of first-hour breastfeeding and 6 months’ exclusive breastfeeding should be amplified.

Similar to the global figures, breastfeeding coverage in Indonesia has been far below the global target. The Basic Health Survey conducted regularly by the Ministry of Health of Indonesia reported a decline in breastfeeding over the past 5 years, falling from 38% in 2013 to 37.2% in 2018 [4,5]. These figures remain far from the national target of 50% in 2019. The good news is that early breastfeeding initiation has risen from 34.5% to 58.2% over the same period [6]. Nevertheless, greater efforts need to be undertaken to improve this figure.

The factors that affect breastfeeding rates are numerous and complex and operate differently in different situations, such as gender inequality, culture, and local norms, as well as the family’s economic situation [3,7]. At the health system level, the role of policy in supporting and regulating breastfeeding promotion in the health delivery setting is crucial, given that the breastfeeding initiation process occurs here [7–9]. Furthermore, the low rates of breastfeeding are attributable in part to a lack of appropriate support by health professionals [10]. In 1991, the World Health Organization (WHO)/United Nations International Children’s Emergency Fund (UNICEF) launched the Baby-Friendly Hospital Initiative (BFHI) [11], based on the Ten Steps to Successful Breastfeeding (TSSB), to increase support for breastfeeding at the healthcare level [7]. A number of studies that evaluated the implementation of TSSB at the hospital level have demonstrated that increased implementation of the TSSB is associated with increased breastfeeding [9,10,12,13]. The studies further suggested that hospitals with comprehensive breastfeeding policies are likely to have better breastfeeding support services and better breastfeeding outcomes [10,12].

Following the international breastfeeding campaign’s agenda, the Indonesian Government has enshrined its commitment to supporting breastfeeding in the form of national regulations. Law Number 36 of 2009 on Health affirms that every child has the right to be breastfed and lays out sanctions for parties who obstruct exclusive breastfeeding [14]. To implement the provisions of Article 129 paragraph (2) of Law Number 36 of 2009 on Health, the Indonesian Government issued Government Regulation Number 33 of 2012 on Exclusive Breastfeeding [15]. Previously, the Ministry of Health of Indonesia had reinforced the exclusive breastfeeding campaign with Ministerial Decree Number 450 of 2004 on Exclusive Breastfeeding [16], which urged health providers to support breastfeeding initiation and exclusive breastfeeding through the implementation of the TSSB. However, none of these health-related regulations have provided systematic guidelines on how health facilities should implement the TSSB, including how to monitor and evaluate its implementation. Interestingly, the guidelines for implementing the TSSB in healthcare facilities were regulated by the Ministry for Women’s Empowerment and Child Protection Regulation Number 3 of 2010. Unfortunately, within the Indonesian constitutional system, the health sector is not directly responsible to the Ministry for Women’s Empowerment and Child Protection and therefore does not have a direct obligation to obey its regulations [17]. At the implementation level, the TSSB policy would require sufficient resources to support information sharing, training, monitoring, and enforcement. With no direct regulation from the health sector authority, there could be limited resources available for the TSSB campaign [18].

With no direct regulation from the health sector authority to support and monitor the implementation of the TSSB in healthcare settings, Indonesia has lacked evidence on the success of its implementation, thereby providing a disincentive for healthcare providers to seriously implement the program. Reports indicate that the implementation of the TSSB and BFHI has long been neglected, although efforts are being made to revitalize them [19]. A published study using Indonesian Demographic and Health Survey data reported an increase in the likelihood of delayed breastfeeding initiation among infants delivered by Cesarean section and in government-owned facilities, where most of the deliveries occurred [20]. This finding clearly highlights the need for implementing the TSSB to support breastfeeding initiation in healthcare facilities in Indonesia. In addition, the BFHI campaign in Indonesia has focused more on hospitals than on other health facilities that organize childbirth assistance, such as public health centers (puskesmas) and private maternity clinics.

With limited reports available on the implementation of the TSSB in Indonesia in hospitals and other healthcare settings, this study aimed to fill the gap by presenting the results of the TSSB implementation survey in healthcare facilities that assist childbirth in 5 cities/districts in East Java, Indonesia.

## Methods

### Study settings

The study was conducted in 5 districts/cities in East Java, namely, Bondowoso, Jember, Probolinggo, Surabaya, and Trenggalek. These areas were selected because of their involvement in the Children Under Two Years Old (*bayi bawah dua tahun* [Baduta]) 2.0 program. The Baduta is a cooperative health program between the Global Alliance for Improved Nutrition (GAIN) and the Ministry of Health of Indonesia aimed at improving nutrition in the first 1000 days of life to prevent stunting. Improving breastfeeding practices is one of Baduta’s main objectives, with a major plan to support the implementation of the TSSB in all healthcare facilities assisting childbirth in 2020.

To implement an appropriate campaign strategy, a baseline survey was conducted to assess the level of TSSB implementation in those 5 areas. The survey was conducted in 242 health facilities (consisting of 22 hospitals, 112 puskesmas, and 108 private maternity clinics) between January and June, 2019, by the Center for Public Health Innovation, Faculty of Medicine, Udayana University.

### Study design, data collection, and analysis

This study adopted a cross-sectional approach aimed at mapping the health facilities in the study areas related to their implementation of the TSSB. The instrument for TSSB implementation was adapted from the BFHI WHO/UNICEF guidelines (Table 1) [21]. The total sample consisted of 372 respondents, including 242 health facility managers (self-appraisal) and 130 postpartum mothers (validation). Interviews with the postpartum mothers were conducted to cross-validate the facility managers’ response regarding several steps of the TSSB. Validation data were only collected on steps 3, 4, 5, 7, 8, and 9 due to the suitability of these items for validation questionnaires.

**Table 1.** Criteria on the Baby-Friendly Hospital Initiative

| Ten Steps to Successful Breastfeeding |  |
| --- | --- |
| Step 1 | Have a written breastfeeding policy that is routinely communicated to all health care staff. (13 items) |
| Step 2 | Train all health care staff in the skills necessary to implement policy. (10 items) |
| Step 3 | Inform all pregnant women about the benefits and management of breastfeeding. (8 items) |
| Step 4 | Facilitate immediate and uninterrupted skin-to-skin contact and support mothers to initiate breastfeeding within an hour of birth. (5 items) |
| Step 5 | Show mothers how to breastfeed and how to maintain lactation even if they should be separated from their infants. (10 items) |
| Step 6 | Give newborn infants no food or drink other than breast milk unless there is a medical indication. (15 items) |
| Step 7 | Rooming-in. (4 items) |
| Step 8 | Breastfeeding on demand. (3 items) |
| Step 9 | No artificial teats or pacifiers to infants. (2 items) |
| Step 10 | Foster the establishment of breastfeeding support groups and refer mothers to them after discharge. (7 items) |

Health facilities were selected using a simple random sampling method, and the selected healthcare facilities were contacted for interviews. The managers interviewed were those responsible for managing the antenatal care (ANC) and childbirth departments or wards. The postpartum mothers from selected healthcare facilities were chosen using incidental sampling on the same day after the manager interviews were completed. Table 2 lists the details on the number of survey participants. Data collection was performed by trained enumerators and retrieved through an e-questionnaire using the Epicollect5 application. Observation-based data collection was also conducted to gather the supporting documents of BFHI implementation at healthcare facilities.

**Table 2.**
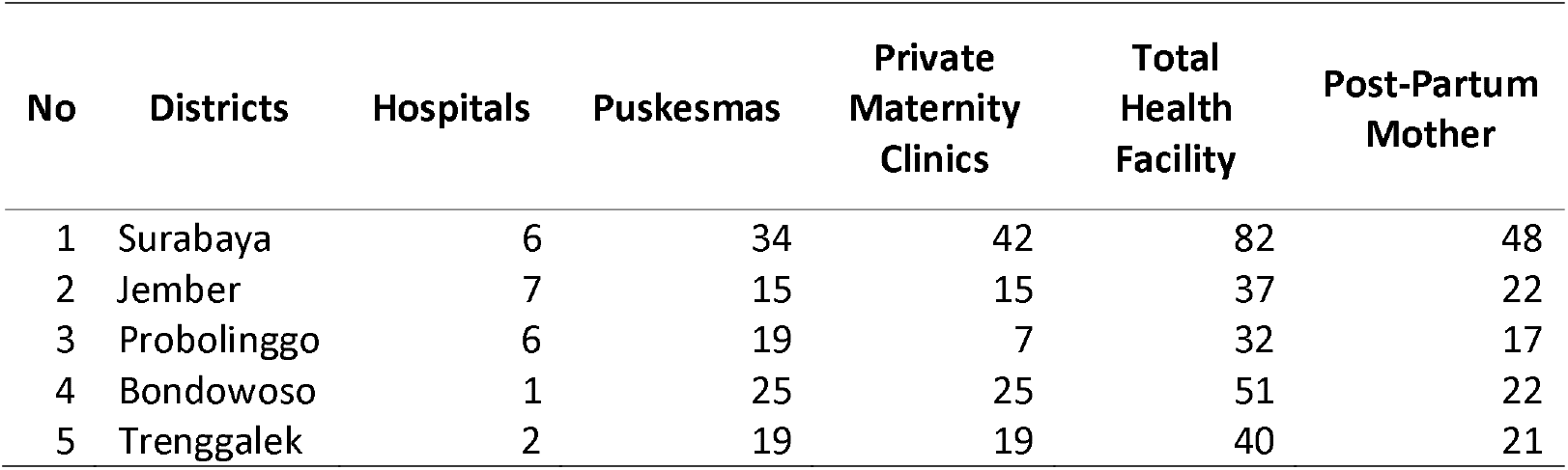
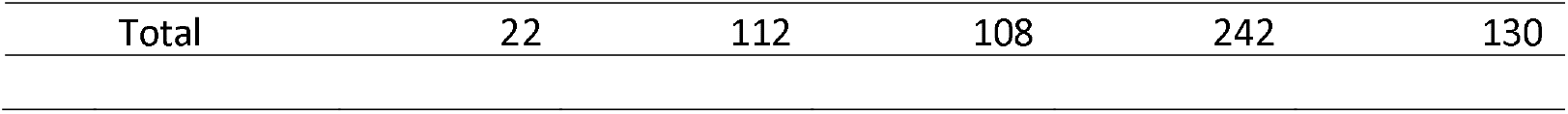
Number of Surveyed Respondents

The data were tabulated on 2 different computers by 2 different researchers (double data entry), and the results were cross-checked to determine data cleaning errors. The data set was processed using 2 different programs (Microsoft Excel and STATA version 12) to identify significant trends in accordance with the research objectives. The TSSB score was 0 or 1 point, attributed to each indicator based on its implementation; a value of 0 denotes that the indicator was not implemented, and a value of 1 denotes that the indicator was implemented. The total score was calculated by adding all the points in each step, then dividing by the maximum points attributed in each step and multiplying by 100, resulting in a score that ranged from 0 to 100 [22]. The self-appraisal and validation results were compared using Two-sample t-tests, and the mean compliance of each facility and district was compared using a One-way ANOVA, with a 5% significance value.

### Ethical considerations

All study participants received information regarding the study implementation from the study’s enumerators and provided signed consent for their participation in the study. The study received ethics approval from the Ethics Committee of the Institute of Research and Community Service, Universitas Katolik Indonesia Atma Jaya, Indonesia (number 0155/III/LPPM-PM.10.05/02/2019).

## Results

Tables 3 and 4 present the socio-demographic characteristics of the healthcare providers and the mothers who were interviewed for validation purposes. From the data analysis, we found various levels of compliance with the TSSB, ranging from 65.47 to 98.09 (mean score of 77.75 on a scale from 0 to 100). The highest compliance was found in step 8 (breastfeeding on demand, 98.09; standard deviation [SD], 7.76]), and the lowest compliance was found in step 7 (rooming-in, 65.47; SD, 25.84). We also found a low level of compliance (<80) in steps 1, 2, 4, 6, and 7. Table 5 presents the comparison between the self-appraisal results (health facility managers) and the validation results (postpartum mothers) in terms of BFHI conformity. The self-appraisal compliance was clearly prone to be higher than the validation result. The validation results showed significantly lower compliance with steps 3, 4, 8, and 9, with a mean (SD) compliance of 78.08 (22.87), 59.81 (26.87), 91.03 (21.48), and 82.69 (32.77), respectively. The largest gap between the self-appraisal and validation results was found in step 4 (skin-to-skin contact).

**Table 3.** Socio-demographic characteristics of health facilities respondents

| Variables | Frequency (%) |
| --- | --- |
| **City/district** |  |
| Bondowoso | 51 (20.90) |
| Jember | 37 (15.16) |
| Probolinggo | 34 (13.93) |
| Surabaya | 82 (33.61) |
| Trenggalek | 40 (16.39) |
| **Health facilities** |  |
| Community health center | 112 (45.90) |
| Private maternity clinics and Private Midwifery Practice | 108 (44.26) |
| Hospital | 24 (9.84) |
| **Age (years)** |  |
| 21-30 | 24 (9.84) |
| 31-40 | 54 (22.13) |
| 41-50 | 118 (48.36) |
| >50 | 48 (19.67) |
| **Sex** |  |
| Male | 20 (8.20) |
| Female | 224 (91.80) |
| **Qualification** |  |
| Public administration | 1 (0.41) |
| Midwife | 190 (77.87) |
| Physician | 26 (10.66) |
| Dentist | 1 (0.41) |
| Nutritionist | 18 (7.38) |
| Public health practitioner | 4 (1.64) |
| Nurse | 2 (0.82) |
| Psychologist | 1 (0.41) |
| Others | 1 (0.41) |
| **Education** |  |
| High school | 1 (0.41) |
| Diploma (3 years) | 68 (27.87) |
| Diploma (4 years) | 128 (52.46) |
| Bachelor's degree | 36 (14.75) |
| Master | 11 (4.51) |
| **Position** |  |
| Head of facilities | 190 (77.87) |
| Head of unit | 31 (12.70) |
| Head of functional staff | 23 (9.43) |
| **Working duration (years)** |  |
| 1-10 | 123 (50.41) |
| 11-20 | 65 (26.64) |
| 21-30 | 44 (18.03) |
| >31 | 12 (4.92) |

**Table 4.** Socio-demographic characteristics of mothers interviewed for validation purposes Variable Frequency (%)

| **Variable** | **Frequency (%)** |
| --- | --- |
| **City or district** |  |
| Bondowoso | 22 (16.92) |
| Jember | 22 (16.92) |
| Probolinggo | 17 (13.08) |
| Surabaya | 48 (36.92) |
| Trenggalek | 21 (16.15) |
| **Age (years)** |  |
| ≤ 20 | 9 (6.92) |
| 21-30 | 82 (63.08) |
| 31-40 | 31 (23.85) |
| 41-50 | 7 (5.38) |
| >50 | 1 (0.77) |
| **Education** |  |
| Primary school | 19 (14.62) |
| Junior high school | 24 (18.46) |
| High school | 56 (43.08) |
| Higher education | 31 (23.85) |
| **Delivery methods** |  |
| Vaginal | 87 (66.92) |
| Caesarian delivery (C-Section) | 43 (33.08) |
| **Health Insurance** |  |
| Yes | 83 (63.85) |
| No | 47 (36.15) |

**Table 5.** Comparison of the compliance with the Ten Steps to Successful Breastfeeding between self-appraisal and validation

| **Steps** | **Self-appraisal** |  | **Validation** |  | **p-value** |
| --- | --- | --- | --- | --- | --- |
|  | **Mean** | **SD** | **Mean** | **SD** |  |
| Step 1 | 66.52 | 25.38 |  |  |  |
| Step 2 | 69.22 | 24.22 |  |  |  |
| Step 3 | 88.83 | 22.24 | 78.08 | 22.87 | ≤0.0001 |
| Step 4 | 66.72 | 25.22 | 59.81 | 26.87 | 0.0141 |
| Step 5 | 82.25 | 17.60 | 81.28 | 26.90 | 0.6744 |
| Step 6 | 79.54 | 15.41 |  |  |  |
| Step 7 | 65.47 | 25.84 | 59.81 | 29.76 | 0.0565 |
| Step 8 | 98.09 | 7.76 | 91.03 | 21.48 | ≤0.0001 |
| Step 9 | 94.67 | 16.11 | 82.69 | 32.77 | ≤0.0001 |
| Step 10 | 89.23 | 15.91 |  |  |  |
| Total Compliance | 77.75 | 11.85 | 74.65 | 16.72 | 0.0385 |

The p-values are derived from Two-sample t-test with equal variance. The level of significance is 5%, SD = Standard Deviation.

The assessment of TSSB compliance also showed certain variations between sites (Table 6). For example, Bondowoso had the lowest compliance (mean score, 72.91; SD, 11.52), whereas Trenggalek had the highest compliance (mean score, 85.78; SD, 11.71). Similar mean compliance scores were found in Probolinggo, Jember, and Surabaya (78.38 [SD, 11.69], 78.45 [SD, 11.89], and 76.27 [SD, 10.27], respectively). In terms of the variations in each step, the majority of districts showed a high level of compliance with step 8 (breastfeeding on demand) and step 9 (no artificial teats or pacifiers given to infants) and the lowest was in step 1 (have a written breastfeeding policy) and step 7 (rooming-in). Almost all steps in the TSSB were significantly different between districts/cities except for step 9 (no artificial teats or pacifiers given to infants) and 10 (establishment of breastfeeding support groups).

**Table 6.** Compliance of the Ten Steps to Successful Breastfeeding by different study areas

| Steps | Bondowoso | Probolinggo | Trenggalek | Jember | Surabaya | p-value |
| --- | --- | --- | --- | --- | --- | --- |
|  | Mean (SD) | Mean (SD) | Mean (SD) | Mean (SD) | Mean (SD) |  |
| Step 1 | 57.92 (30.89) | 64.93 (27.58) | 69.23 (23.69) | 64.66 (20.78) | 72.05 (22.07) | 0.0310 |
| Step 2 | 51.96 (23.24) | 75.88 (24.38) | 90.00 (13.39) | 72.43 (20.87) | 65.61 (21.61) | ≤0.0001 |
| Step 3 | 90.68 (22.20) | 95.22 (8.71) | 88.44 (23.91) | 94.93 (11.58) | 82.46 (27.13) | 0.0120 |
| Step 4 | 63.92 (10.59) | 64.12 (27.75) | 77.00 (20.03) | 69.73 (20.34) | 63.17 (32.73) | 0.0428 |
| Step 5 | 80.00 (12.49) | 81.18 (18.38) | 91.50 (13.11) | 82.43 (20.46) | 79.51 (19.30) | 0.0066 |
| Step 6 | 76.86 (11.57) | 76.86 (15.84) | 86.33 (10.34) | 78.92 (14.45) | 79.27 (18.76) | 0.0324 |
| Step 7 | 55.39 (15.25) | 72.06 (21.11) | 87.5 (22.64) | 66.89 (24.33) | 57.62 (27.96) | ≤0.0001 |
| Step 8 | 100.00 | 100.00 | 98.33 (7.36) | 93.69 (13.23) | 97.96 (8.03) | 0.0016 |
| Step 9 | 96.08 (13.58) | 92.64 (17.97) | 98.75 (7.91) | 95.95 (13.83) | 92.07 (19.98) | 0.2105 |
| Step 10 | 89.91 (12.55) | 87.39 (22.99) | 94.27 (19.08) | 88.42 (14.99) | 87.46 (12.37) | 0.2226 |
| Total compliance | 72.91 (11.52) | 78.38 (11.69) | 85.78 (11.71) | 78.45 (11.89) | 76.27 (10.27) | ≤0.0001 |

The p-values are from one-way ANOVA. The level of significance is 5%, SD = Standard Deviation

The mean compliance score of the health facilities with all the BFHI criteria also varied significantly (Table 7). In puskesmas, for instance, the mean compliance score was 78.21 (SD, 11.07). Similar results were found in private maternity clinics (76.61; SD, 12.55). Compared with other facilities, the highest mean compliance score was for hospitals (80.74; SD, 11.96). Variations in each step were also found among the healthcare facilities. Most of the healthcare facilities had high compliance (>90) with steps 8 and 9, and the lowest compliance was with steps 1 and 7. Only steps 2, 4, 7, and 10 showed significant differences between the type of health facilities.

**Table 7.** Compliance of the Ten Steps to Successful Breastfeeding by different health care facilities

| Steps | Community |  | Clinics and Private |  | Hospital |  | p-value |
| --- | --- | --- | --- | --- | --- | --- | --- |
|  | Health Centre |  | Midwifery Practice |  |  |  |  |
|  | mean | SD | mean | SD | Mean | SD |  |
| Step 1 | 67.45 | 22.81 | 65.59 | 28.36 | 66.35 | 23.33 | 0.8650 |
| Step 2 | 74.20 | 22.77 | 62.59 | 25.22 | 75.83 | 19.09 | 0.0006 |
| Step 3 | 87.61 | 25.08 | 89.93 | 20.18 | 89.58 | 16.76 | 0.7321 |
| Step 4 | 62.32 | 27.47 | 67.96 | 22.86 | 81.67 | 17.61 | 0.0021 |
| Step 5 | 79.91 | 17.27 | 83.52 | 17.94 | 87.5 | 16.48 | 0.0962 |
| Step 6 | 80.71 | 13.84 | 77.41 | 17.41 | 83.61 | 11.29 | 0.1108 |
| Step 7 | 62.5 | 26.63 | 64.35 | 24.46 | 84.38 | 20.61 | 0.0006 |
| Step 8 | 97.62 | 8.62 | 98.15 | 7.67 | 100.00 | 0.00 | 0.3945 |
| Step 9 | 95.08 | 16.38 | 94.44 | 15.79 | 93.75 | 16.89 | 0.9167 |
| Step 10 | 92.60 | 12.56 | 89.15 | 15.36 | 73.81 | 22.55 | 0.0000 |
| Total Compliance | 78.21 | 11.07 | 76.61 | 12.55 | 80.74 | 11.96 | 0.2614 |

The p-values are from one-way ANOVA. The level of significance is 5%, SD = Standard Deviation

## Discussion

To the best of our knowledge, this is the first study in the published literature to show compliance with the TSSB of the BFHI across different health facilities. This is also the first study to report compliance with the TSSB in Indonesian settings, including large-scale sites and facilities. Most of the relevant studies in the published literature have reported only on TSSB compliance in hospital settings [23–25], and almost all of the studies found in the literature assessing the impact of compliance on breastfeeding outcomes were conducted in hospital settings [12,26,27]. This situation could be due to the use of the term “hospital” in the WHO/UNICEF TSSB campaign, which provided a loophole in many middle-low income countries, given that a large proportion of ANC and deliveries occur outside hospitals, particularly in healthcare centers and private clinics.

Regarding compliance with the TSSB criteria, steps 1, 2, 4, and 7 scored low compliance (<80), with the lowest compliance found for steps 1 (policy) and 7 (rooming-in). The low compliance level for step 1 occurred across cities and healthcare facilities, indicating the need to strengthen efforts to promote TSSB implementation at a policy level. A study conducted in Oregon found that hospitals with comprehensive breastfeeding policies were likely to have better breastfeeding support services and better breastfeeding outcomes [12]. Policy changes, including better policy communication, are needed to promote the implementation of the next steps of the TSSB [12,28].

The low level of compliance with step 1 (policy) was followed by a relatively low level of compliance with step 2 (staff training). An inadequate policy for implementing the TSSB resulted in the absence of a legal framework, guidance, and commitment to improving the staff’s understanding and related skills [28,29]. Poor compliance with step 2 affects the coverage of exclusive breastfeeding. A study performed in Canada revealed that the training of hospital nurses could significantly increase the coverage of exclusive breastfeeding, from 31% to 54% [30].

Furthermore, the low level of compliance found in steps 4 (skin to skin contact) and 7 (rooming-in) could be related to the lack of health facility infrastructure and the number of medical staff, as well as their capacity to support immediate skin-to-skin contact and rooming-in practices [31,32]. We found that the low compliance level was significantly different across healthcare facilities, in which puskesmas showed the poorest compliance, followed by private clinics. Hospitals had the highest compliance. Other factors that influenced the decision for rooming-in were staff experience with rooming-in practices, breastfeeding counseling training, parity, and delivery methods [33].

Overall, the self-appraisal results were prone to higher values compared with the validation data, in which steps 3 (information), 4 (skin to skin contact), 8 (breastfeeding on demand), and 9 (no artificial teats or pacifiers given to infants) had significant differences between self-appraisal and validation. A possible reason for this is the fact that healthcare facility leaders are prone to overestimating the factual data to maintain their institution’s image. It might therefore be difficult for these respondents to be objective in their self-appraisal of the respective health facility’s performance [22]. However, a higher score in the validation data could well be caused by the external position of the mother and her ability to provide an accurate overview of the health facility’s services. These considerations indicate that the self-appraisal results were unlikely to provide a true reflection of each health facility’s compliance with the BFHI criteria. The largest gap between the self-appraisal and validation results was found in step 4. Our result was in line with a study performed in Croatia, which found that compliance with step 4 was also <50% [34]. In Indonesia, the percentage of Cesarean-sections was relatively high (9.8% in 2013), which inhibits compliance with step 4. This was evidenced by American studies that revealed that giving birth by Cesarean section represents a significant barrier to early breastfeeding initiation [35,36].

The highest level of compliance across the 4 districts/cities and also across health facilities was in step 8 (encouraging breastfeeding on demand), which shows the effort by health facilities to support exclusive breastfeeding. The results also revealed that certain steps showed high conformity, namely steps 3, 5, 9, and 10 and indicates the institution’s attempts to apply the TSSB, despite the high rate of violation with step 1 of having written policy [37].

## Limitations of the study

This study has 3 major limitations. First, the cross-sectional data collection method represents short-term data and cannot assess longitudinal TSSB conformity [38]. Second, the data representation and analysis represent a weakness because, of the 244 healthcare facility managers, only 130 (53.28%) participated in the validation; 46.72% of the healthcare facilities were not verified. Lastly, validation by the postpartum mothers on the health facilities’ compliance was only performed on steps 3, 4, 5, 7, 8, and 9 due to the suitability of these items for the questionnaire. The rest of the steps were not subject to validation. Future studies should address these concerns.

## Conclusions

This study concluded that a low level of TSSB compliance was particularly related to steps 1, 2, 4, and 7. We recommend robust steps to encourage policy makers and managers of healthcare facilities to formulate effective and appropriate policies to raise institutional compliance with the TSSB of the BFHI. Greater effort is needed to facilitate puskesmas and private clinics in implementing the TSSB due to their significant involvement in providing ANC and deliveries in Indonesia. Furthermore, there is an imperative need for providing refresher training for staff.

## Data Availability

All data produced in the present study are available upon reasonable request to the authors

## Acknowledgments

We would like to thank the Health Office of East Java Province and the Health Office of each study site for helping us with the administrative aspects of the study. We offer our gratitude to all of the health facility managers and mothers who participated in the survey. Lastly, we would like to thank GAIN Indonesia for providing us with resources to conduct the study and write the report.

## References

1. UNICEF, WHO. Increasing commitment to breastfeeding through funding and improved policies and programmes: Global breastfeeding scorecard 2019. Geneva: UNICEF, WHO; 2019.

2. Oddy WH. Breastfeeding protects against illness and infection in infants and children: a review of the evidence. Breastfeed Rev. Nursing Mothers’ Association of Australia; 2001;9:11.

3. Horta BL, Bahl R, Martinés JC, Victora CG, Organization WH. Evidence on the long-term effects of breastfeeding: systematic review and meta-analyses. World Health Organization; 2007;

4. Ministry of Health of the Republic of Indonesia. Basic Health Research (Riskesdas) 2013. Jakarta; 2013.

5. Ministry of Health of the Republic of Indonesia. Basic Health Research (Riskesdas) 2018. Jakarta; 2018.

6. Ministry of Health of the Republic of Indonesia. Strategic Plan of the Ministry of Health of the Republic of the Indonesia for 2015-2019. Jakarta; 2015.

7. World Health Organization (WHO). Evidence for the ten steps to successful breastfeeding. Geneva: World Health Organization; 1998.

8. Bhandari N, Kabir IAKM, Salam MA. Mainstreaming nutrition into maternal and child health programmes: Scaling up of exclusive breastfeeding. Matern Child Nutr. 2008;4:5–23.

9. Li CM, Li R, Ashley CG, Smiley JM, Cohen JH, Dee DL. Associations of hospital staff training and policies with early breastfeeding practices. J Hum Lact. 2014;30:88–96.

10. Gomez-Pomar E, Blubaugh R. The Baby Friendly Hospital Initiative and the ten steps for successful breastfeeding. a critical review of the literature. J Perinatol [Internet]. Springer US; 2018;38:623–32. Available from: http://dx.doi.org/10.1038/s41372-018-0068-0

11. WHO. Indicators for assessing breastfeeding practices. Geneva: World Health Organization Geneva; 1991.

12. Rosenberg KD, Stull JD, Adler MR, Kasehagen LJ, Crivelli-Kovach A. Impact of hospital policies on breastfeeding outcomes. Breastfeed Med. Mary Ann Liebert, Inc. 140 Huguenot Street, 3rd Floor New Rochelle, NY 10801 …; 2008;3:110–6.

13. Sinha B, Chowdhury R, Sankar MJ, Martines J, Taneja S, Mazumder S, et al. Interventions to improve breastfeeding outcomes: a systematic review and meta-analysis. Acta Paediatr. Wiley Online Library; 2015;104:114–34.

14. Government Republic of Indonesia. Law on Health (Law No. 36/2009). 36 0f 2009 Indonesia; 2009.

15. Government Republic of Indonesia. Government Regulation Number 33 of 2012 on Exclusive Breast Feeding. 33 tahun 2012 Indonesia; 2012.

16. Ministry of Health of Indonesia. Minister of Health Decree No. 450 of 2004 on Exclusive Breastfeeding in Indonesia. 450/menkes/SK/IV/2004 Jakarta, Indonesia; 2004.

17. Ministry of Women Empowerment and Child Protection. Ministry of Women Empowerment and Child Protection No. 3 of 2010 on the STSB implementation. Indonesia; 2010.

18. Soekarjo D, Zehner E. Legislation should support optimal breastfeeding practices and access to low-cost, high-quality complementary foods: Indonesia provides a case study. Matern Child Nutr. 2011;7:112–22.

19. Trisnantoro L, Soemantri S, Singgih B, Pritasari K, Mulati E, Agung FH, et al. Reducing child mortality in Indonesia. SciELO Public Health; 2010.

20. Titaley CR, Loh PC, Prasetyo S, Ariawan I, Shankar AH. Socio-economic factors and use of maternal health services are associated with delayed initiation and non-exclusive breastfeeding in Indonesia: Secondary analysis of Indonesia Demographic and Health Surveys 2002/2003 and 2007. Asia Pac J Clin Nutr. 2014;23:91–104.

21. World Health Organization/ United Nations Children’s. Baby-Friendly Hospital Initiative Revised Updated and Expanded for Integrated Care Section 3 Breastfeeding Promotion and Support in a Baby-Friendly Hospital 20-hour Course for Maternity Staff. Geneva; 2009.

22. Araújo RG, de Fonseca V de M, de Oliveira MIC, Ramos EG. External evaluation and self-monitoring of the Baby-friendly Hospital Initiative’s maternity hospitals in Brazil. Int Breastfeed J. International Breastfeeding Journal; 2019;14:1–9.

23. Haiek LN. Measuring compliance with the Baby-Friendly Hospital Initiative. Public Health Nutr. 2012;15:894–905.

24. Zakarija-Grković I, Boban M, Janković S, Ćuže A, Burmaz T. Compliance with who/unicef bfhi standards in croatia after implementation of the bfhi. J Hum Lact. 2018;34:106–15.

25. Grizzard TA, Bartick M, Nikolov M, Griffin BA, Lee KG. Policies and practices related to breastfeeding in Massachusetts: Hospital implementation of the Ten Steps to Successful Breastfeeding. Matern Child Health J. 2006;10:247–63.

26. Hawkins SS, Stern AD, Baum CF, Gillman MW. Compliance with the Baby-Friendly Hospital Initiative and impact on breastfeeding rates. Arch Dis Childhood-Fetal Neonatal Ed. BMJ Publishing Group; 2014;99:F138–43.

27. Nickel NC, Labbok MH, Hudgens MG, Daniels JL. The extent that noncompliance with the ten steps to successful breastfeeding influences breastfeeding duration. J Hum Lact. 2013;29:59–70.

28. Okolo SN, Ogbonna C. Knowledge, attitude and practice of health workers in Keffi local government hospitals regarding Baby-Friendly Hospital Initiative (BFHI) practices. Eur J Clin Nutr. Nature Publishing Group; 2002;56:438–41.

29. Balogun OO, Dagvadorj A, Yourkavitch J, Da Silva Lopes K, Suto M, Takemoto Y, et al. Health Facility Staff Training for Improving Breastfeeding Outcome: A Systematic Review for Step 2 of the Baby-Friendly Hospital Initiative. Breastfeed Med [Internet]. 2017;12:537–46. Available from: http://www.liebertpub.com/doi/10.1089/bfm.2017.0040

30. Martens PJ. Does Breastfeeding Education Affect Nursing Staff Beliefs, Exclusive Breastfeeding Rates, and Baby-Friendly Hospital Initiative Compliance? The Experience of a Small, Rural Canadian Hospital. J Hum Lact. SAGE Publications Inc.; 2000;16:309–18.

31. Taylor EC, Nickel NC, Labbok MH. Implementing the Ten Steps for Successful Breastfeeding in Hospitals Serving Low-Wealth Patients. Am J Public Health. 2012;102:2262–8.

32. Kakrani VA, Rathod Waghela HK, Mammulwar MS, Bhawalkar JS. Awareness about “Ten Steps for Successful Breastfeeding” among Medical and Nursing Students. Int J Prev Med. Isfahan University of Medical Sciences(IUMS); 2015;2015-May.

33. Hakala M, Kaakinen P, Kääriäinen M, Bloigu R, Hannula L, Elo S. Implementation of Step 7 of the Baby-Friendly Hospital Initiative (BFHI) in Finland: Rooming-in according to Mothers and Maternity-ward Staff. Eur J Midwifery. E.U. European Publishing; 2018;2.

34. Grguric J, Zakarija-Grkovic I, Pavičić Bošnjak A, Stanojevic M. A Multifaceted Approach to Revitalizing the Baby-Friendly Hospital Initiative in Croatia. J Hum Lact. SAGE Publications Inc.; 2016;32:568–73.

35. Crenshaw JT, Cadwell K, Brimdyr K, Widström AM, Svensson K, Champion JD, et al. Use of a Video-Ethnographic Intervention (PRECESS Immersion Method) to Improve Skin-to-Skin Care and Breastfeeding Rates. Breastfeed Med. 2012;7:69–78.

36. Weddig J, Baker SS, Auld G. Perspectives of Hospital-Based Nurses on Breastfeeding Initiation Best Practices. JOGNN - J Obstet Gynecol Neonatal Nurs. Blackwell Publishing Ltd; 2011;40:166–78.

37. Fikawati S SA. Study on Implementation and Exclusive Breastfeeding Policy and Early Breastfeeding Initiation in Indonesia. Makara Kesehat. 2010;14:17–24.

38. Kovach AC. A 5-year follow-up study of hospital breastfeeding policies in the Philadelphia area: a comparison with the ten steps. J Hum Lact. Sage Publications Sage CA: Thousand Oaks, CA; 2002;18:144–54.

